# The Persistence of Neutralising Antibodies up to 11 months after SARS-CoV-2 Infection in the Southern Region of New Zealand

**DOI:** 10.1101/2021.10.26.21265501

**Authors:** Reuben McGregor, Alyson Craigie, Susan Jack, Arlo Upton, Nicole J. Moreland, James Ussher

## Abstract

During the first wave of SARS-CoV-2 infection in New Zealand a cohort of 78 PCR-confirmed COVID-19 cases was recruited in the Southern District Health Board region. Here we report on this unique cohort nearly 1-year after infection. There was no known community transmission in the region over the study period due to New Zealand’s elimination status at the time, nor had any participants received a COVID-19 vaccine. In the absence of re-exposure, antibody reactivity to the viral spike protein, as well as neutralising antibodies to both the ancestral strain and the delta variant remained relatively stable between 8 and 11 months post-infection. This suggests long-lived antibody responses can be generated from a single natural infection event. However, given the risks of serious disease associated with SARS-CoV-2 infection, vaccination is still strongly recommended.

---

Severe acute respiratory syndrome coronavirus 2 (SARS-CoV-2), which causes Coronavirus Disease 2019 (COVID-19), was first detected in New Zealand (NZ) in February 2020 (1). Following this initial introduction, and a community outbreak of 1154 confirmed cases, NZ successfully eliminated the virus in the community. With the exception of several isolated border incursions and short lockdowns in Auckland, the country remained largely COVID free until the current outbreak of the Delta SARS-CoV-2 variant, which began in August 2021.

The emergence of novel viral variants of concern (VoC), such as Delta, combined with reports of gradual waning of antibodies over extended timeframes (2) highlights a need for ongoing studies tracking immune responses following natural infections and vaccination. Particularly since the initial waves of infections, and current licenced vaccines, are based on the ancestral SARS-CoV-2 strain rather than the Delta variant, which is now dominating infections globally. Here we present a follow up serological assessment of PCR confirmed cases nearly 1-year post-infection, including levels of neutralising antibodies to alpha, beta and delta VoC.

During the first wave of infection in NZ a cohort of PCR-confirmed COVID-19 cases was recruited in the Southern District Health Board (SDHB) region (3). We have previously reported on antibody dynamics in this cohort, alongside participants from other cohorts, up to 8-months post-infection (4). This showed that antibody (IgG) responses to the viral spike protein and neutralising antibodies were relatively stable over this 8-month period compared with antibodies to the nucleocapsid protein. This persistence of spike specific antibodies compared with the rapid decay of nucleocapsid antibodies has since been widely demonstrated, with the latter now being utilised as a marker of recent infection (5).

The original SDHB cohort comprised n=78 PCR confirmed cases infected between 11/03/2020 and 5/04/2020, with up to three serum samples collected post-symptom onset (Figure 1, clear circles and Table 1). Of these, 30 participants donated further samples at later time points, represented as red circles in Figure 1. As there were no successive community outbreaks in SDHB during the study timeframe, nor had any participants received a COVID-19 vaccine, the immune responses observed likely represent a single exposure event tracked over the time course. Median days post symptom onset for this additional timepoint was 302 days (Table 1). Samples were assayed for antibodies to both nucleocapsid (Abbott Architect SARS-CoV-2 IgG assay, Figure 2a)) and spike proteins (Abbott Alinity SARS-CoV-2 IgG II Quant assay, Figure 2b). We have previously reported 99.7% specificity for the nucleocapsid assay using 300 pre-pandemic ANS samples (3). The same procedure was followed for the recently released Spike Alinty IgG assay for this study, for a calculated specificity of 100% (0/100 above the 50 AU/mL cut-off).

**Table 1.** Cohort demographics.

|  | Total | New |
| --- | --- | --- |
| Participants, *n* (samples, *n*) | 78 (172) | 30 (37) |
| Sex, *n* (M/F) | 31/47 | 9/21 |
| Age (year) |  |  |
| Median | 51.5 | 52 |
| Range | 17-81 | 27-81 |
| DPSO (days) |  |  |
| Median | 158 | 302 |
| Range | 80-344 | 235-344 |

**Figure 1.**
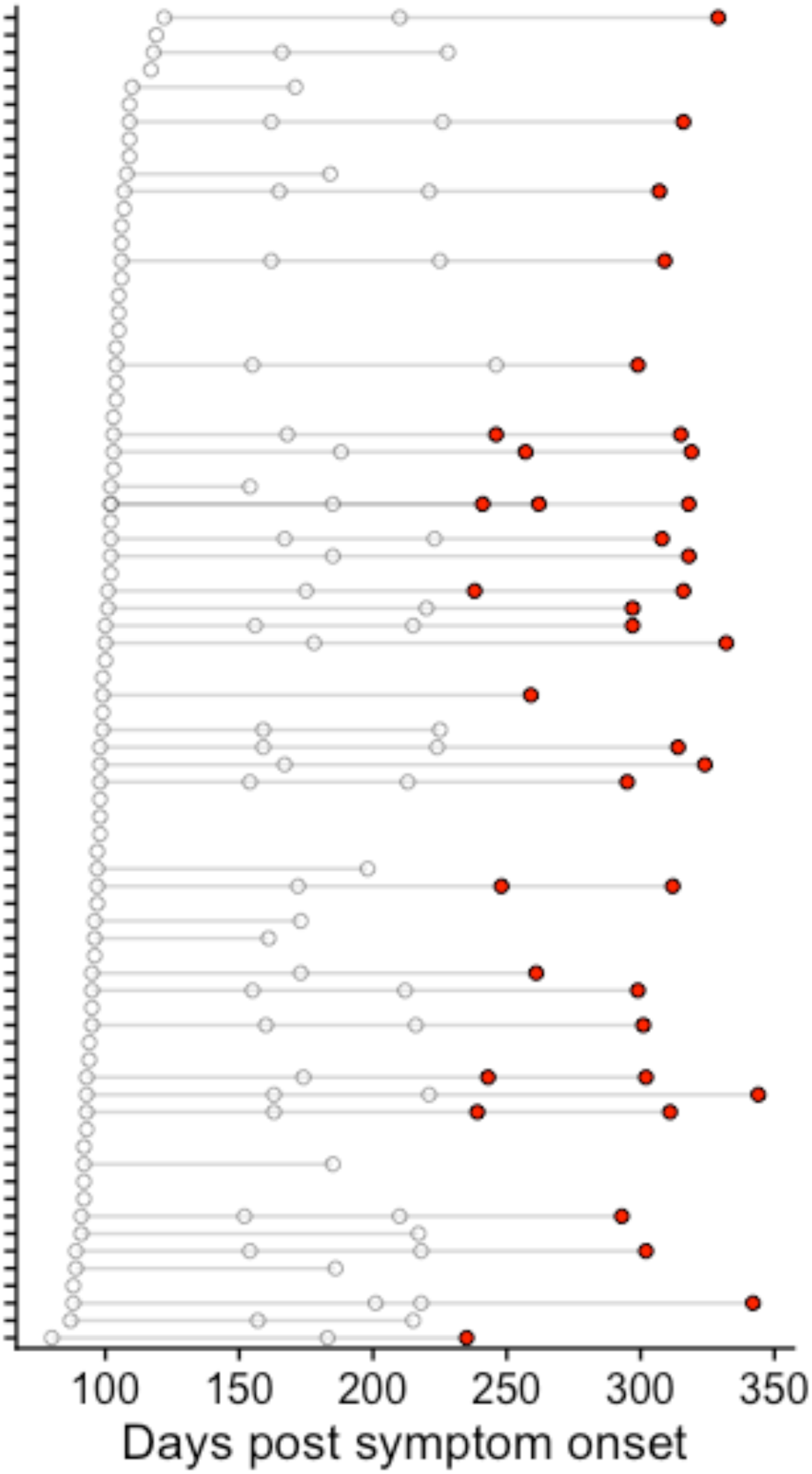
Cohort summary. Individual participants are ordered by earliest donation days post onset of symptoms, with temporal samples from the same individuals connected by grey lines. Participants donated one to four timepoints over the study period. New donations are indicated by red circles with ealier donations inidcated by unfilled circles.

**Figure 2.**
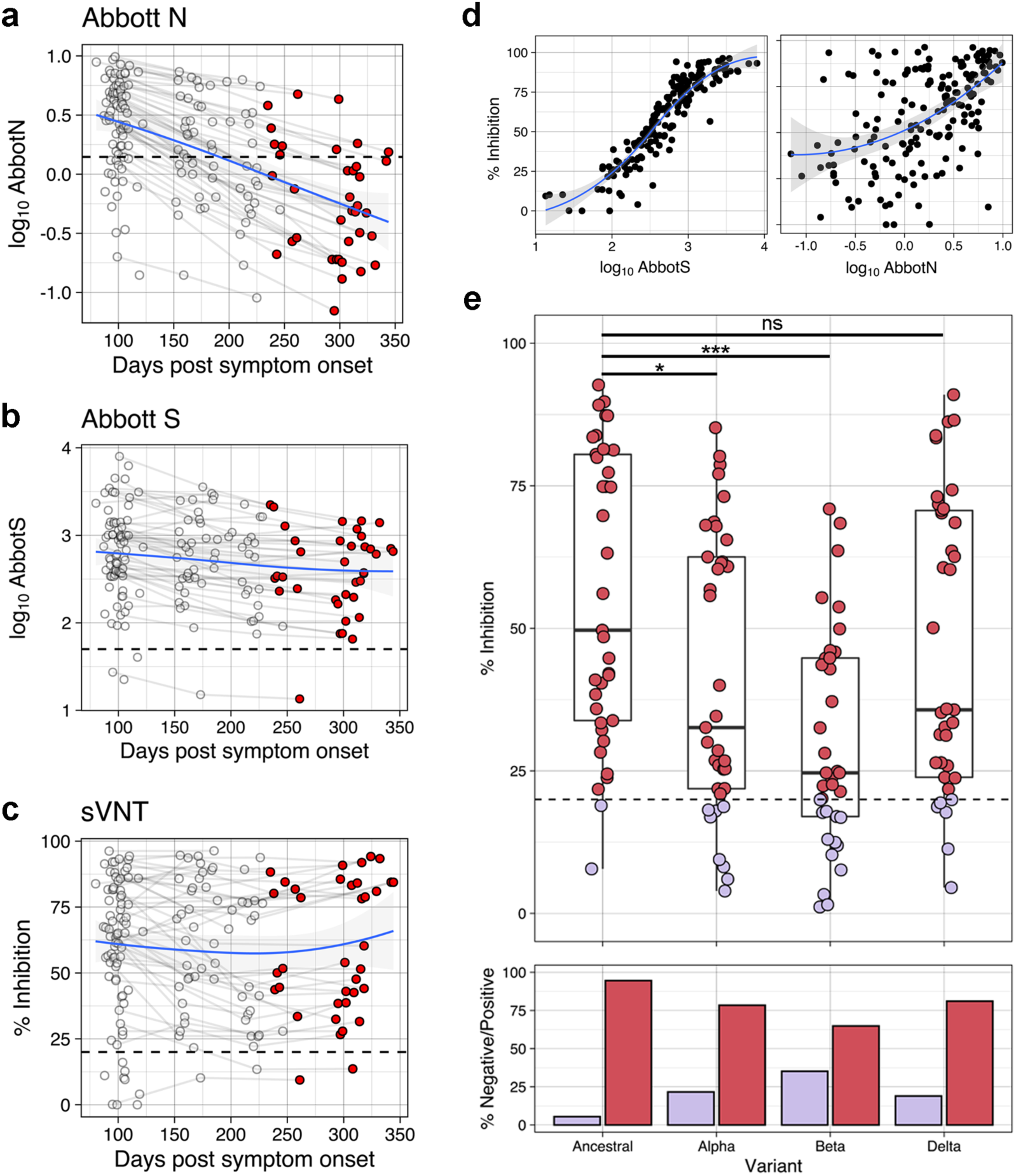
Antibody responses following SARS-CoV-2 infection over time. Antibody responses targeting Nucleocapsid (N) protein **(a)**, Spike (S) protein **(b)** as well as neutralising antibodies **(c)** over time. New donations are indicated by red circles with previously reported donations inidcated by unfilled circles (n=172). **(d)** Correlation between S protein antibodies (left) and N protein antibodies (right) vs neutralising antibodies. LOESS regression (blue line) was used to visualise general trends in antibody levels, and standard error of regression is indicated by grey shaded area (n=172). **(e)** Neutralising antibody responses to variants of concern including only the latest donations (represented by red circles in a-c) (top). Kruskal-Wallis test showed a significant difference (p<0.001) with follow up Wilcoxen test Holm adjusted p-values indicated by stars, *p<0.05, ***p<0.001 (n=37). Percentage of samples below (purple) and above (pink) the assay cut-off (bottom). Dashed horizontal lines represent respective test cut-off’s throughout.

Neutralising antibodies were measured using a surrogate Viral Neutralisation test (sVNT), based on the receptor binding domain of the spike protein (cPass™ SARS-CoV-2 Neutralization Antibody Detection Kit, GenScript). This domain contains >90% of neutralising antibody epitopes - that is regions that block the entry of the virus into host cells via the hACE-2 receptor (6). Specificity was previously determined to be 100% using the 300 pre-pandemic ANS, as well as an additional 113 pre-pandemic samples (7).

Recent analyses suggest that the level of neutralising antibodies is an important component of a correlate of protection (8). There is now intense effort on understanding how sequence changes within the receptor binding domain in VoC might impact on neutralising antibody activity, and protection from re-infection (6). In this study neutralising antibodies were assessed to alpha (B.1.1.7, United Kingdom), beta (B.1.351, South Africa) and delta (B.1.617.2, India) VoC using an adapted sVNT assay that incorporates receptor binding domains corresponding to the sequence for each of these variants (9).

With the inclusion of later time-points, the relative stability of spike antibodies (Figure 2b), compared with nucleocapsid antibodies (Figure 2a) has been confirmed in this cohort. Indeed, nucleocapsid antibodies continue to decline rapidly between 8-11 months, with 27/37 (72.97%) below the cut-off at this later time-point. This contrasts with spike antibodies where 36/37 (97.30%) remained positive at the later time-point. Similarly, neutralising antibodies to the ancestral SARS-CoV-2 virus showed no decline at this later timepoint (Figure 2c), with 35/37 (94.59%) remaining positive. This trend was also reflected in the strong correlation between spike and neutralising antibodies, which was not observed with nucleocapsid antibodies (Figure 2d).

To assess the neutralisation capacity of sera against VoC generated by a single natural exposure approximately 300 days prior, the sVNT with alpha, beta and delta was performed following standard protocols (4,9). There was a highly significant reduction in neutralisation capacity to the beta variant compared with ancestral strain (median inhibition 24.7% versus 49.7%, p<0.0001) and a significant, but less marked, reduction to the alpha variant (32.6%, p<0.05). In contrast, there was a non-significant reduction to the delta variant (35.7%, p=0.117) (Figure 2e). This trend was mirrored in the proportion of samples below the assay cut-off (<20% inhibition), with beta showing the highest proportion of negative samples (13/37, 35.1%), compared with alpha (8/37, 21.6%) and delta (7/37, 18.9%) (Figure 2e).

These data are in keeping with observations internationally, where the beta variant is thought to evade humoral immunity compared to the alpha and delta variants (2). This is partly driven by the E484K mutation in the beta receptor binding domain that interferes with antibodies generated in response to ancestral strain sequences (10). In contrast, the delta variant does not contain this critical immune escape mutation but is instead associated with increased transmissibility due to gains in replication efficiency (2,11). The persistent levels of neutralising antibodies to the delta variant following ancestral strain infection observed bodes well for vaccination with ancestral spike protein sequences such as Comirnaty (Pfizer/BioNTech), being administered in NZ.

In conclusion, this study provides novel insight from a unique setting in the Southern region of NZ where the probability of SARS-CoV-2 re-exposure has been extremely unlikely. A single exposure has been shown to generate a neutralising antibody response, including to the delta VoC, that persists for at least 11 months. However, given the risks of serious disease associated with SARS-CoV-2 infection, vaccination is still strongly recommended.

## Data Availability

All data produced in the present study are available upon reasonable request to the authors

## Acknowledgments

We thank the team behind the Southern COVID-19 Serology Study for facilitating sample collection, delivery, and processing for the SARS-CoV-2 serological testing. We thank GenScript for providing the VoC for the sVNT testing kits and technical advice. Sample collection, processing, and Abbott Architect consumables were funded by Southern Community Laboratories. This work was also funded in part by the School of Medicine Foundation (University of Auckland).

